# Factors Associated With Postoperative Complications In Adult Patients Undergoing Cataract Surgery At KCMC Hospital 2023-2024

**DOI:** 10.1101/2025.07.02.25330702

**Authors:** Jeremiah Msika Kaduga, Einot Matayan, Andrew Makupa

## Abstract

**Objective:** This study aims to assess the incidence, types, and associated risk factors of postoperative complications following cataract surgery at Kilimanjaro Christian Medical Centre (KCMC) from 2023 to 2024

**Background:** Cataracts remain a leading cause of blindness globally, with disproportionate burdens in low-resource settings. Despite advancements in surgical techniques like phacoemulsification (PHACO), extracapsular cataract extraction (ECCE) remains widely used in sub-Saharan Africa. Limited data exist on postoperative complications and comparative outcomes in Tanzania.

**Methods:** A prospective cohort study was conducted at Kilimanjaro Christian Medical Centre (August 2023–July 2024) involving 308 adults undergoing ECCE or PHACO. Participants were followed for six months postoperatively. Primary outcomes included visual acuity (VA) and complications (e.g., corneal edema, raised intraocular pressure [IOP]). Multivariable logistic regression identified risk factors for poor outcomes.

**Results:** The cohort (median age: 68 years; 51.6% male) underwent ECCE (65.3%) or PHACO (34.7%). PHACO demonstrated superior outcomes: patients undergoing ECCE had 2.5-fold higher odds of poor VA (adjusted OR: 2.4, 95% CI: 1.21–5.01, p=0.013) and 34% higher complication risk (adjusted RR: 0.66, 95% CI: 0.50–0.88, p=0.004). Complications occurred in 58.8%, primarily corneal edema (40.6%) and posterior capsular opacity (38%). Specialist-performed surgeries reduced complications by 62% versus trainees (p<0.001).

**Conclusion:** PHACO outperformed ECCE in visual outcomes and safety, with specialist expertise further reducing risks. Findings support prioritizing PHACO adoption, enhancing surgical training, and standardizing postoperative care in resource-limited settings. Multicenter randomized trials are needed to validate these results.

## Introduction

Cataracts remain a leading cause of blindness globally, with cases of cataract-related blindness rising from 12.3 million in 1990 to 20 million by (1) (2).They contribute to approximately 40% of global blindness (3), and cataract-related disability-adjusted life years (DALYs) increased by over 90% between 1990 and 2019 (4). The burden is especially high in low-income regions, with sub-Saharan Africa reporting cataract-related blindness rates ranging from 21% to 67% (5). In Tanzania, cataracts account for nearly 48% of bilateral visual impairment (6).

Cataract surgery is the only definitive treatment, offering substantial improvements in visual acuity and quality of life (Davis, 2016; Rossi *et al.*, 2021). It is one of the most frequently performed surgeries worldwide, with more than 26 million procedures annually (10). The most commonly used surgical techniques include extracapsular cataract extraction (ECCE), manual small-incision cataract surgery (MSICS), and phacoemulsification (11,12). Advances in surgical methods—particularly phacoemulsification—have led to faster recovery, better outcomes, and reduced complication rates (13).

MSICS is considered a cost-effective alternative to phacoemulsification, especially in low-resource settings, due to its shorter learning curve, low cost, and comparable outcomes (14). Despite improvements, cataract surgery can still lead to postoperative complications such as corneal edema, increased intraocular pressure (IOP), posterior capsular opacification (PCO), cystoid macular edema, uveitis, and retinal detachment (15,16).

Global studies have reported varying complication rates. In Ethiopia, early complications included raised IOP (42.3%), striate keratopathy (48.7%), and corneal edema (36.6%) (17).Complications can also result from systemic comorbidities such as diabetes, hypertension, and renal disease (18).

The risk of complications also varies with surgical technique, surgeon experience, and patient comorbidities (Stein et al, 2013). Good outcomes are associated with younger age, male sex, and absence of inflammation, while poor visual outcomes correlate with baseline poor vision and intraoperative complications (20).

Despite global data, there is a lack of region-specific studies in Tanzania addressing postoperative complications and their associated risk factors.

This study aims to assess the incidence, types, and associated risk factors of postoperative complications following cataract surgery at Kilimanjaro Christian Medical Centre (KCMC) from 2023 to 2024.

## Materials and Methods

This was a prospective cohort study conducted at the Eye Department of Kilimanjaro Christian Medical Centre (KCMC), a tertiary referral hospital in northern Tanzania. The study took place from August 20, 2023, to July 1, 2024. Participants were recruited between August 20 and December 31, 2023, and were followed at two week then monthly for six months postoperatively to monitor for complications. At each visit, patients underwent a full ophthalmic examination, including standardized intraocular pressure (IOP) measurement.

Ethical approval was obtained from the Kilimanjaro Christian Medical University College Research and Ethical Committee (Ref: PG.88/2023), and all patients gave written informed consent. The study adhered to the Declaration of Helsinki principles throughout its implementation.

The study population included all adult patients (aged 18 and above) undergoing either extracapsular cataract extraction (ECCE) or phacoemulsification (PHACO) at KCMC who consented to participate. Exclusion criteria were pre-existing corneal diseases (e.g., corneal opacities, dystrophies, keratoconus, pterygium, or synechiae), and any cognitive impairment that could interfere with participation or follow-up.

Participants were selected using purposive sampling to include individuals meeting the inclusion criteria and scheduled for surgery within the designated timeframe. This approach ensured the selection of clinically relevant participants who reflected a range of demographic and ocular conditions.

Data were collected using structured questionnaires capturing demographic and clinical information, along with standardized ophthalmic examination results. Pre- and postoperative assessments included visual acuity using a Snellen chart, IOP measurement with an iCare tonometer, fundus examination using indirect ophthalmoscopy with a 90D lens, and corneal endothelial cell density measured via specular microscopy (preoperatively and six weeks postoperatively).

The primary outcome variables were postoperative complications and visual acuity outcomes. Independent variables included demographic characteristics (age, sex, residence, and occupation), clinical history (e.g., comorbidities), and type of cataract, surgical technique, and surgeon experience.

Data analysis was performed using STATA version 17. Descriptive statistics were presented using means, medians, and proportions. Categorical variables were compared using chi-square tests. Logistic regression analysis was employed to identify factors associated with postoperative complications. Univariate analysis produced crude odds ratios (COR), and variables with p < 0.05 were included in multivariable models to calculate adjusted odds ratios (AOR). A p-value of < 0.05 was considered statistically significant.

## Results

Table 1 below represents the social demographic of study participants. The study involved 308 participants with 59.4% falling between 61 and 80 years, 27.6% aged 60 or younger, and 13% older than 80 years. The median age was 68, with an interquartile range of 60 to 75. Regarding gender, the sample was evenly split with 51.6% male and 48.4% female. As for their residence, 68.2% were from Kilimanjaro, while the remaining 31.8% lived outside the Kilimanjaro region.

**Table 1.** Clinical characteristics of study participants.

| **Variable** | **Frequency** | **Percentage** |
| --- | --- | --- |
| **Age of Participant** |  |  |
| ≤60 | 85 | 27.6 |
| 61-80 | 183 | 59.4 |
| >80 | 40 | 13 |
| *Median (IQR)* | *68(60-75)* |  |
| **Sex of the participants** |  |  |
| Male | 159 | 51.6 |
| Female | 149 | 48.4 |
| **Residence** |  |  |
| Out of Kilimanjaro | 98 | 31.8 |
| Kilimanjaro | 210 | 68.2 |
| **Pre-operative ECD** |  |  |
| Normal | 113 | 36.7 |
| Abnormal | 195 | 63.3 |
| *Median(IQR)* |  |  |
| **Pre-operative IOP (mmHg)** |  |  |
| Normal | 297 | 96.4 |
| Abnormal | 11 | 3.6 |
| *Mean (SD)* | *14.6(±2.83)* |  |
| **Best corrected VA** |  |  |
| Poor | 177 | 57.5 |
| Good | 131 | 42.5 |
| **Surgeon experience** |  |  |
| AMO | 15 | 4.9 |
| Resident | 141 | 45.8 |
| Specialist | 152 | 49.4 |

The majority of participants 75% did not have a systemic chronic disease, while 25% had systematic chronic disease. ECCE was the most common type of cataract surgery performed, accounting for 65.3% of the cases, while those who underwent PHACO were 34.7%. The majority of participants had mature cataracts 80.8%, with the remaining participants having hyper-mature cataracts 19.2%.

Also, before the pre-operative surgery, 63.3% of the participants had abnormal ECD, and the median IOP was 14.6 mmHg (±2.83 SD). During the study, 96.4% of the participants had normal IOP levels, while 3.6% had abnormal levels. The BCVA was poor for 57.5% of the participants and good for 42.5%. The surgeries were performed by a mix of medical professionals — 4.9% by AMO, 45.8% by residents, and 49.4% by specialists.

**“In Table 2”**, the factors associated with poor VA two weeks after operative surgery are detailed.

**Table 2.** Factors associated with poor VA two weeks post-operative.

|  | VA |  | COR (95%CI) | P-value | AOR (95%CI) | P-value |
| --- | --- | --- | --- | --- | --- | --- |
|  | Poor | Good |  |  |  |  |
| **Age** |  |  |  |  |  |  |
| 39-60 | 10(16.7) | 73(29.4) | 1 |  | 1 |  |
| 61-80 | 39(65.0) | 145(58.5) | 0.51(0.24-1.08) | 0.078 | 0.96(0.33-2.76) | 0.933 |
| 80-92 | 11(18.3) | 30(12.1) | 0.37(0.14-0.97) | **0.044** | 0.69(0.20-2.33) | 0.546 |
| **Gender** |  |  |  |  |  |  |
| Male | 33(55.0) | 126(50.8) | 0.85(0.48-1.49) | 0.560 | 0.75(0.42-1.35) | 0.343 |
| Female | 27(45.0) | 122(49.2) | 1 |  | 1 |  |
| **Cataract surgery performed** |  |  |  |  |  |  |
| ECCE | 48(80.0) | 156(62.9) | 2.36(1.19-4.67) | **0.014** | 2.46(1.21-5.01) | **0.013\*** |
| PHACO | 12(20.0) | 92(37.1) | 1 |  | 1 |  |
| **IOL decentration** |  |  |  |  |  |  |
| Yes | 2(3.3) | 1(0.4) | 0.81(0.46-1.44) | 0.476 | 0.10(0.01-1.21) | 0.070 |
| No | 58(96.7) | 247(99.6) | 1 |  | 1 |  |
| **PCO** |  |  |  |  |  |  |
| Yes | 25(41.7) | 91(36.7) | 0.12(0.01-1.32) | 0.082 | 0.75(0.41-1.37) | 0.348 |
| No | 35(58.3) | 157(63.3) | 1 |  | 1 |  |
| **Cystoid Macular Edema** |  |  |  |  |  |  |
| Yes | 1(1.7) | 1(0.4) | 0.24(0.01-3.87) | 0.314 | 0.21(0.01-3.47) | 0.276 |
| No | 59(98.3) | 247(99.6) | 1 |  | 1 |  |

In multivariate logistic regression, those who underwent ECCE cataract surgery were 2.5 times more likely to have poor VA compared to PHACO, a statistically significant (COR=2.4, 95% CI 1.21-5.01, P-value=0.013).

The overall incidence of post-cataract complications was 58.8% (181/308). The distribution of types of post-cataract complications according to their type is shown in “**Figure 1”** below, which summarizes the complications observed among post-cataract surgery patients whereby cornea edema125(40.6%), raised IOP 122(39.6%), posterior capsular opacity 117(38%), and fibrin reaction 81(26.3%) were the most common observed complications among the study participants.

**Figure 1.**
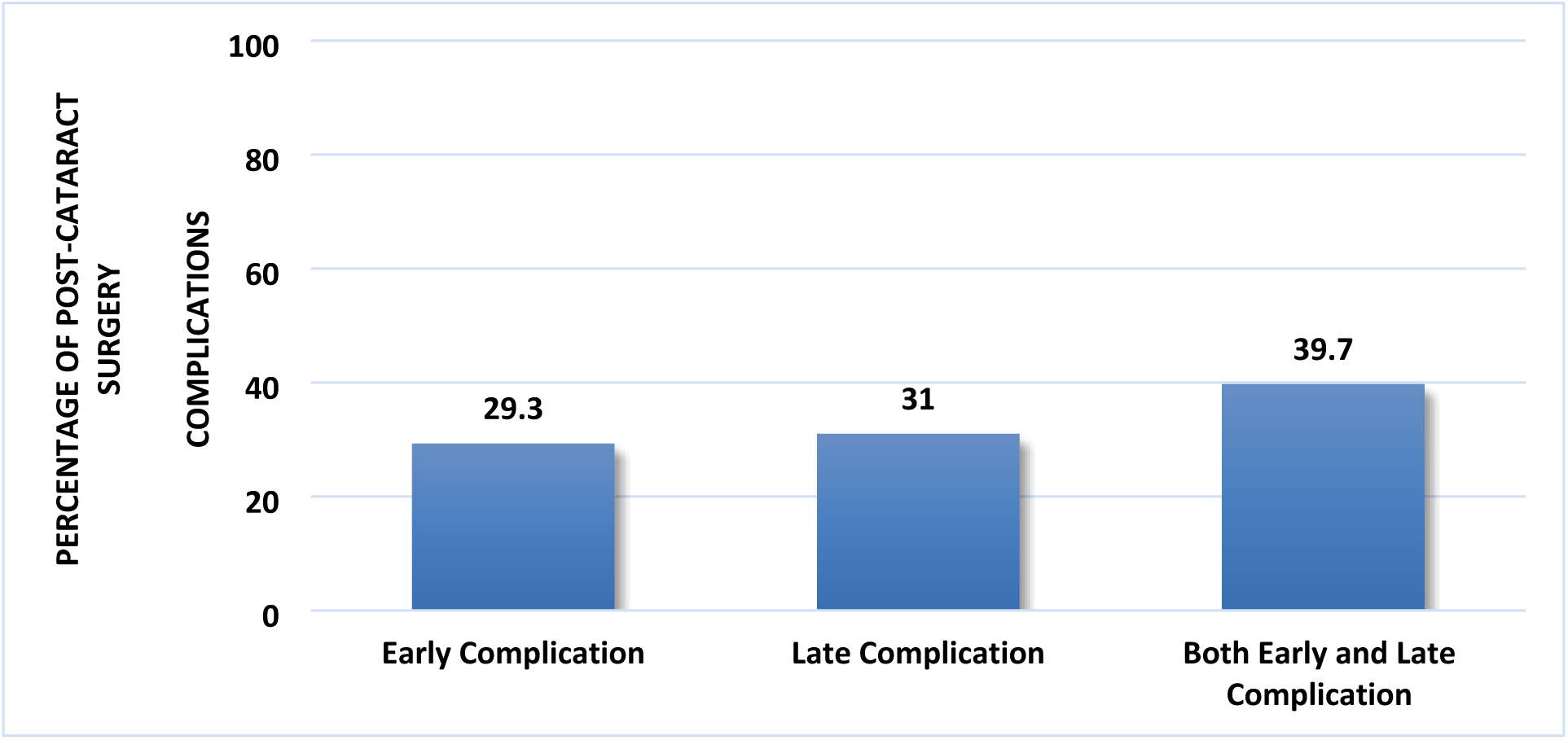
Percentage of type complications after post-cataract surgery (N=181)

The study reported both early and late complications following cataract surgery see ***“Table 3***”. Early complications included corneal edema in 32% of patients and raised IOP in 5%. Additionally, fibrin reaction occurred in 6.1% of cases, and hyphema was observed in 1.6%. Late complications were primarily posterior capsular opacity, affecting 33% of patients. Other late complications included astigmatism in 7.7% of cases, IOL decentralization in 2.8%, and cystoid macular edema in 1.1%.

**Table 3.** Complications of post-cataract surgery (N=308)

| Variable | Frequency | Percentage |
| --- | --- | --- |
| **Early complications** | 55 | 30.4 |
| Cornea Edema | 32 | 17.6 |
| Fibrin reaction | 11 | 6.1 |
| Hyphema | 3 | 1.6 |
| Raised IOP | 9 | 5 |
| **Late complications** | 54 | 29.8 |
| Posterior capsular Opacity | 33 | 18.2 |
| IOL decentralization | 5 | 2.8 |
| Cystoid Macular Edema | 2 | 1.1 |
| Astigmatism | 14 | 7.7 |

**“Table 4**”, summarizes factors associated with associated with post-cataract surgery complications among study participants. In crude analysis type of cataract surgery performed, pre-operative IOP and surgeon experience were factors that were statistically significantly associated with post-cataract surgery complications.

**Table 4.** Factors associated with post-cataract surgery complications.

| Variable | CRR (95%CI) | P-value | ARR (95%CI) | P-value |
| --- | --- | --- | --- | --- |
| **Age of Participant** |  |  |  |  |
| ≤60 | Ref |  |  |  |
| >60 | 1.03(0.84-1.28) | 0.735 |  |  |
| **Sex of the participants** |  |  |  |  |
| Male | Ref |  |  |  |
| Female | 0.78(0.49-1.24) |  |  |  |
| **Systemic Chronic disease** |  |  |  |  |
| No | Ref |  |  |  |
| Yes | 1.11(0.91-1.35) | 0.312 |  |  |
| **Cataract surgery performed** |  |  |  |  |
| ECCE | Ref |  | Ref |  |
| PHACO | 0.70(0.56-0.88) | 0.003 | 0.66(0.50-0.88) | 0.004 |
| **Type of Cataract** |  |  |  |  |
| Mature | Ref |  |  |  |
| Hypermature | 1.10(0.88-1.36) | 0.403 |  |  |
| **Pre-operative ECD** |  |  |  |  |
| Normal | Ref |  |  |  |
| Abnormal | 1.04(0.86-1.26) | 0.652 |  |  |
| **Pre-operative IOP** |  |  |  |  |
| Normal | Ref |  | Ref |  |
| Abnormal | 1.40(1.04-1.88) | 0.026 | 1.28(0.99-1.66) | 0.058 |
| **Surgeon experience** |  |  |  |  |
| Trainee | Ref |  |  |  |
| Specialist | 0.41(0.31-0.56) | <0.001 | 0.38(0.28-0.53) | <0.001 |

Compared to patients who underwent ECCE, patients who underwent PHACO had 30% significantly less risk of having post-cataract surgery complications (CRR=0.70; 95%CI: 0.56-0.88; P-value= 0.003), While compared to patients who had normal preoperative IOP, patients who had abnormal Preoperative IOP had 40% significantly high risk of having post-cataract surgery complications (CRR=1.40; 95%CI:1.04-1.88; P-value= 0.026). Compared to Trainee, the risk of having post-cataract surgery complications was 59% lower (CRR=0.41; 95%CI: 0.31-0.56; P-value<0.001) for surgeries performed by specialists.

In adjusted analysis, the type of cataract surgery performed and surgeon experience were factors that were statistically significantly associated with post-cataract surgery complications. After adjusting for pre-operative IOP and surgeon experience, compared to patients who underwent ECCE, patients who underwent PHACO had 34% significantly less risk of having post-cataract surgery complications (ARR=0.66; 95%CI: 0.50-0.88; P-value= 0.004), While compared to Trainee, the risk of having post-cataract surgery complications was 62% lower for surgeries performed by specialists (CRR=0.38; 95%CI: 0.28-0.53; P-value<0.001) adjusting for the type of cataract surgery performed and pre-operative IOP.

## Discussion

The types of cataract surgeries performed (ECCE and PHACO) and their outcomes were a central focus. ECCE was the more commonly performed procedure, but it was associated with a higher likelihood of poor VA compared to PHACO. This finding was consistent in both univariate and multivariate analyses, suggesting that PHACO might be a more effective technique for improving postoperative VA. The presence of corneal edema and raised intraocular pressure (IOP) were significant predictors of poor VA, indicating the importance of managing these complications to improve surgical outcomes.

The study also reported a high incidence of post-cataract surgery complications (58.8%) with corneal edema, raised IOP, and posterior capsular opacity being the most common. The findings underscore the need for careful postoperative monitoring and management to minimize these complications. Surgeries performed by specialists had a significantly lower risk of complications compared to those performed by trainees, emphasizing the role of surgeon experience in improving surgical outcomes.

This study findings were similar to findings of previous studies done in Ghana and Ethiopia in which the prevalence of post cataract surgery complications was 48.6% and 55% (21); (17) respectively. These findings can be due to the reason that quality of postoperative care, surgical techniques and study design and methodology contribute to postoperative complications. However, KCMC being a Zonal Hospital with well-equipped in terms of advanced techniques and technology together with enough skillful man power, could be a reason for a relatively high Cataract Surgical Rate (CSR) that performed annually and hence this can adversely lead to drastically increase in postoperative complications.

In this study early, late and complications were observed following cataract surgery. In early complications corneal edema was the leading complication after cataract surgery (17.6%) followed by and fibrin reaction (6.1%), raised IOP (5%) and hyphema (1.6%). In late complications posterior capsular opacity was the leading complication (18.2%) followed by astigmatism (7.7%), IOL decentralization (2.8%), and cystoid macular edema in 1.1%.

These results were similar to the study done by Nampradit and Kongsapin 2021,Lin et al 2021 (22,23). Also, Long-term complications were similar in PCO 8.71%. Both studies indicate that Hyphema is the most common early complication after Operative cataract surgery and PCO the latest complication after cataract operative surgery.

In two weeks, the factors associated with poor VA for those who underwent ECCE cataract surgery were 2.5 times more likely to have poor VA compared to PHACO this could be explained by PHACO involve small incision compared to ECCE..

The finding is similar to our studies done by Thanigasalam, Reddy and Zaki (24).

The results were significantly different from the study done by Markos,Tamrat and Asferaw (17). Where Age-related macular degeneration and preoperative astigmatism were significantly associated with poor postoperative visual outcome. The difference could time interval of VA after surgery where factors were observed from 6 to 8 weeks.

Also our finding is different from study done in Eastern region Ghana where the factors associated with poor VA were postsurgical complications, biometry, comorbid, and complications during surgery (21).

Surgical technique were statistically significantly associated with post-cataract surgery complications. Patients who underwent ECCE, patients who underwent PHACO had 34% significantly less risk of having post-cataract surgery complications.

While compared to Trainee, the risk of having post-cataract surgery complications was 62% lower for surgeries performed by specialists, this explain the importance of surgical experience of specialist compared to trainee.

The results were similar to the study done Thanigasalam, Reddy,and Zaki, (24).

Hover the findings differed from the study in United States, where associated factors for complication were divorced status, never marriage, diabetes with ophthalmic manifestations, traumatic cataract, previous ocular surgery, and old age (25).

The following are some limitation of the study, the study was conducted at a single center and it was hospital based, which may limit the generalizability of the results to other settings or populations and the absence of randomization in assigning surgical techniques could affect the comparability of the ECCE and PHACO groups. In conclusion this study demonstrated PHACO’s superiority over ECCE in cataract surgery, with better visual outcomes and fewer complications. Specialist surgeons achieved significantly better results than trainees, emphasizing surgical skill’s importance. The findings support adopting PHACO while highlighting needs for technique refinement and better postoperative care

## Data Availability

All relevant data are within the manuscript

## Acknowledgement

I would like to express my deep and sincere gratitude to my research supervisor, Dr Einoti Matayan and Dr. Andrew Makupa as a Co-Supervisor for providing valuable guidance throughout this project. I would like to thank them for being flexible, motivated and sincerity, they have deeply inspired me. It was a great privilege and honor to work and study under their invaluable guidance. I am extremely grateful for what they have offered me from the beginning of this project throughout the process of compiling of this document. This expertise and input were incredibly instrumental in shaping the final outcome.

I am also extremely grateful to all the specialists, in the eye department who provided their day-to-day support, as well as my fellow residents, for their continuous encouragement and assistance.

Last but certainly not least, I want to express my deepest gratitude to my lovely wife for her dedicatedly prayers for me and endless social support for the whole period of this research work. I am also expressing my thanks to my Daughters, Grace, Gladness and Glory for their valuable encouragement and prayers. Their presence and endless encouragement to me have been a driving force behind my determination to accomplish this project

